# A Bayesian model for quantifying genomic variant evidence sufficiency in Mendelian disease

**DOI:** 10.64898/2025.12.02.25341503

**Authors:** Quant Group, Ali Saadat, Shweta Pipaliya, Dylan Lawless

## Abstract

**Summary:** Clinical genomic interpretation depends on heterogeneous evidence checks that vary across institutions and pipelines. This variation prevents evidence from being compared or verified in a reliable way. We address this logistical barrier by introducing a universal, verifiable layer that evaluates the completeness of available scientific evidence independently of upstream analytic systems. Quantitative Evidence Sufficiency (Quant ES) summarises, for each variant, how much verifiable evidence is available using a binary matrix derived from registered rule sets. A closed form Beta Binomial model produces a single evidence sufficiency estimate with a credible interval and genome wide percentile. This separates evidence availability from provider-specific logic and enables consistent interpretation and reuse of results across institutions.

**Implementation:** QuantBayes implements the method as compiled C binaries and an R package. It requires no access to proprietary algorithms and scales to millions of variants, allowing integration into existing diagnostic and research workflows without modification to upstream systems.

**Availability:** The QuantBayes software releases for macOS and Linux are at https://doi.org/10.5281/zenodo.17919369. The QuantBayes R package is on the Comprehensive R Archive Network (CRAN) https://doi.org/10.32614/CRAN.package.quantbayes. All releases are under the MIT licence.

**Graphical abstract:** Genomic analyses produce candidate variants, but the completeness of their supporting evidence is not directly comparable across institutions. Quant Evidence Sufficiency operates on a binary evidence matrix to summarise how much verifiable scientific evidence is available for each variant. This separates analysis outputs from interpretation and delivers an interpretable, uncertainty-calibrated measure of credibility that can be shared and reused without exposing provider-specific methods or intellectual property.

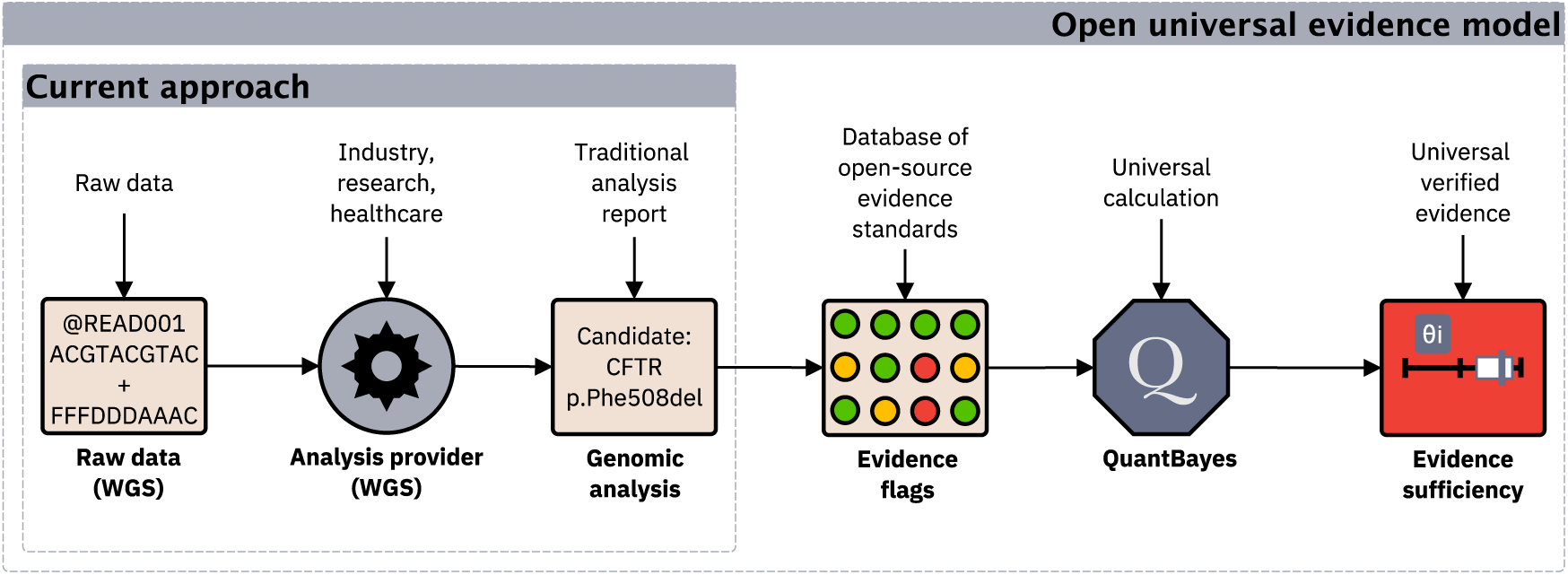

## 1 Introduction

Clinical variant interpretation relies on analysis that include biological plausibility, inheritance consistency, population frequency, and phenotype fit (1–6). These assessments are implemented differently across pipelines, so the evidential basis for a candidate cannot be reliably compared or verified across institutions (7). Structured specifications such as the qualifying variant framework (8) and the American College of Medical Genetics and Genomics (ACMG) criteria (1; 2) provide practical ways to generate verifiable evidence flags. These include checks such as confirming inheritance consistency or excluding variants that are too common to explain a rare disorder. These rules can be evaluated for any pipeline result without disclosure of proprietary logic.

Nevertheless, the field lacks a statistical measure of how complete and trustworthy the resulting verifiable evidence is for a specific candidate. Manual inspection and heuristic scoring do not quantify uncertainty and do not place candidates within the wider evidential landscape.

This gap reflects a broader logistical challenge. Institutions encode and interpret evidence differently, yet clinical decision making requires a shared, reproducible basis for assessing how well supported each candidate is. A framework is needed that operates solely on verifiable evidence, remains independent of pipeline internals, and quantifies evidence completeness in a transparent and comparable manner.

Here we introduce the Quantitative Evidence Sufficiency model (Quant ES) and its implementation in the QuantBayes software for quantifying the sufficiency of verifiable scientific evidence supporting individual genomic variants. The framework is grounded in Bayesian principles (9; 10). Existing tools primarily estimate the likelihood that a variant is causal, while the completeness and reliability of the underlying scientific evidence are rarely measured directly (11). Quant ES addresses this gap by providing an independent standardised evidence layer that complements pathogenicity assessment and supports transparent comparison and reuse of results across clinical, research, and commercial settings.

## 2 Methods

### 2.1 Evidence matrix

Genomic variant calling, annotation, and interpretation are implemented heterogeneously across institutions, limiting direct comparability of variant reports (1–6; 11–14). To demonstrate a verifiable and comparable input, we briefly summarise an externally defined normative standard Qualifying Evidence Matrix for verifiable evidence (SGA-QEM-1.0) (7; 8). This specification is independent of Quant ES and encodes rule-based evidence derived from subject-level data and reference databases into standardised evidence flags, which are subsequently interpreted as evidence present or missing (examples in **Box S1–S2** and **Table S1**). Quant ES operates solely on the resulting binary evidence matrix, irrespective of how the flags are generated. For each variant *i*, the interpretation rule set (7; 8) produces a binary vector of length *m*, where

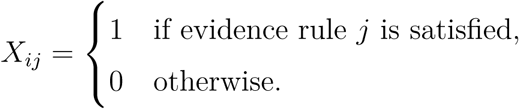

The total number of satisfied rules is 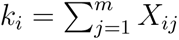, and the empirical proportion of satisfied evidence is *p̂_i_* = *k_i_*/*m*. This quantity summarises the amount of verifiable evidence supporting variant *i*.

Rules are defined so that a TRUE flag always indicates a contradiction or weakening signal and is encoded as *X_ij_* = 0. A FALSE flag indicates that no contradiction is observed and is encoded as *X_ij_* = 1. Missing or unavailable information is also encoded as *X_ij_* = 0 and never contributes evidence. For example, in **Table S1**, a TRUE flag for parental genotype availability indicates that the genotype is unavailable and contributes no evidence (*X_ij_* = 0), whereas a FALSE flag indicates that the genotype is available and contributes evidence (*X_ij_* = 1).

### Bayesian evidence sufficiency model

We define the evidence sufficiency parameter *θ_i_* as the probability that a randomly selected evidence rule is satisfied for variant *i*. The model assumes *k_i_*| *θ_i_* ∼ Binomial(*m, θ_i_*)*, θ_i_* ∼ Beta(1, 1). By conjugacy, the posterior distribution is *θ_i_* | *k_i_*∼ Beta(1 + *k_i_,* 1 + *m* − *k_i_*). The posterior mean and Credible Interval (CrI) (10; 15) provide an uncertainty-calibrated measure of evidence completeness for each variant (9; 16).

The use of a Beta–Binomial model follows directly from the structure of the evidence. Under the adopted standard, each variant is assessed against a fixed set of *m* evidence rules, each of which is either satisfied or not. The observed evidence count *k_i_* therefore represents the number of successes in *m* Bernoulli trials (one per evidence rule) with a common success probability. Modelling *k_i_* with a binomial likelihood captures this process directly, without introducing assumptions about rule weighting, dependence, or pathogenic effect, and reflects the fact that evidence availability is always incomplete rather than evidence being absent.

### Global posterior distribution

To contextualise variant-level evidence, we analyse each *θ_i_* relative to the empirical mixture of all posterior distributions across the genome (17). Monte Carlo draws from each *θ_i_* yield a global distribution from which we estimate genome-wide summary statistics and CrI. For a set of candidate variants, we compute the percentile of each *θ_i_* within this distribution as percentile(*i*) = Pr (*θ_j_* ≤ *θ_i_*), with *j* indexing all evaluated variants. This provides a genome-wide reference for interpreting evidence sufficiency.

### Validation study on a public WGS trio

We evaluated the framework using Whole Genome Sequencing (WGS) from the public Genome in a Bottle (GIAB) family trio (HG005–HG007, PRJNA200694, GRCh38 v4.2.1) (18). All processing parameters were defined in an external qualifying variant specification (8). Quality control and study filters were applied to illustrate a typical clinical WGS pipeline. Site-level thresholds on QUAL and INFO/DP were applied using BCFtools, complemented by per-sample thresholds on FORMAT/DP and FORMAT/GQ with exclusion of missing genotypes.

Composite filters required either all trio samples or at least one sample to pass the criteria. The filtered trio Variant Call Format (VCF) was analysed with Exomiser (default configuration) using the family .ped file and without Human Phenotype Ontology (HPO) terms (19). Exomiser outputs were then processed with the registered qualifying variant evidence standard (7; 8) flag rule set to generate raw per-variant evidence flags (TRUE, FALSE, or NA). A second registered interpretation rule set converted these raw flags into the binary evidence matrix required by Quant ES, with values indicating evidence supporting (1) or missing (0). All rule sets used in this workflow are provided in YAML Ain’t Markup Language (YAML) or JavaScript Object Notation (JSON) format (8). Quant ES was then run on the binary evidence matrix using default settings to generate per-variant verifiable evidence *θ*, 95% CrIs, and genome-wide evidence summaries, implemented with QuantBayes software.

## 3 Implementation

### QuantBayes software

QuantBayes implements the Quant ES model as a lightweight, pipeline-independent tool. It is distributed as precompiled C binaries for macOS and Linux and as a cross platform R package with identical statistical behaviour (20; 21). All implementations operate directly on a binary evidence matrix containing values in {0, 1, NA} and require no access to genotype data, pipeline internals, or proprietary algorithms. The expected input format is illustrated in **Figure 1 (A)**. The standalone C binary runs without installation and is suitable for automated High Performance Computer (HPC) workflows. It produces a per-variant summary including the number of satisfied evidence rules *k_i_*, the number of evaluated rules *m*, the posterior mean of *θ_i_*, its 95% CrI, and the percentile within the genome wide distribution, together with a global summary of the posterior mixture. Optional outputs include a concise human-readable report and structured JSON representations for integration into relational or graph based systems such as Electronic Health Record (EHR) platforms or Resource Description Framework (RDF) stores (22). Per-variant outputs are shown in **Figure 1 (B)**, with an overview of computed quantities in **Figure S2**. All implementations use the same closed form Beta updates and yield identical results up to floating point precision.

**Figure 1:**
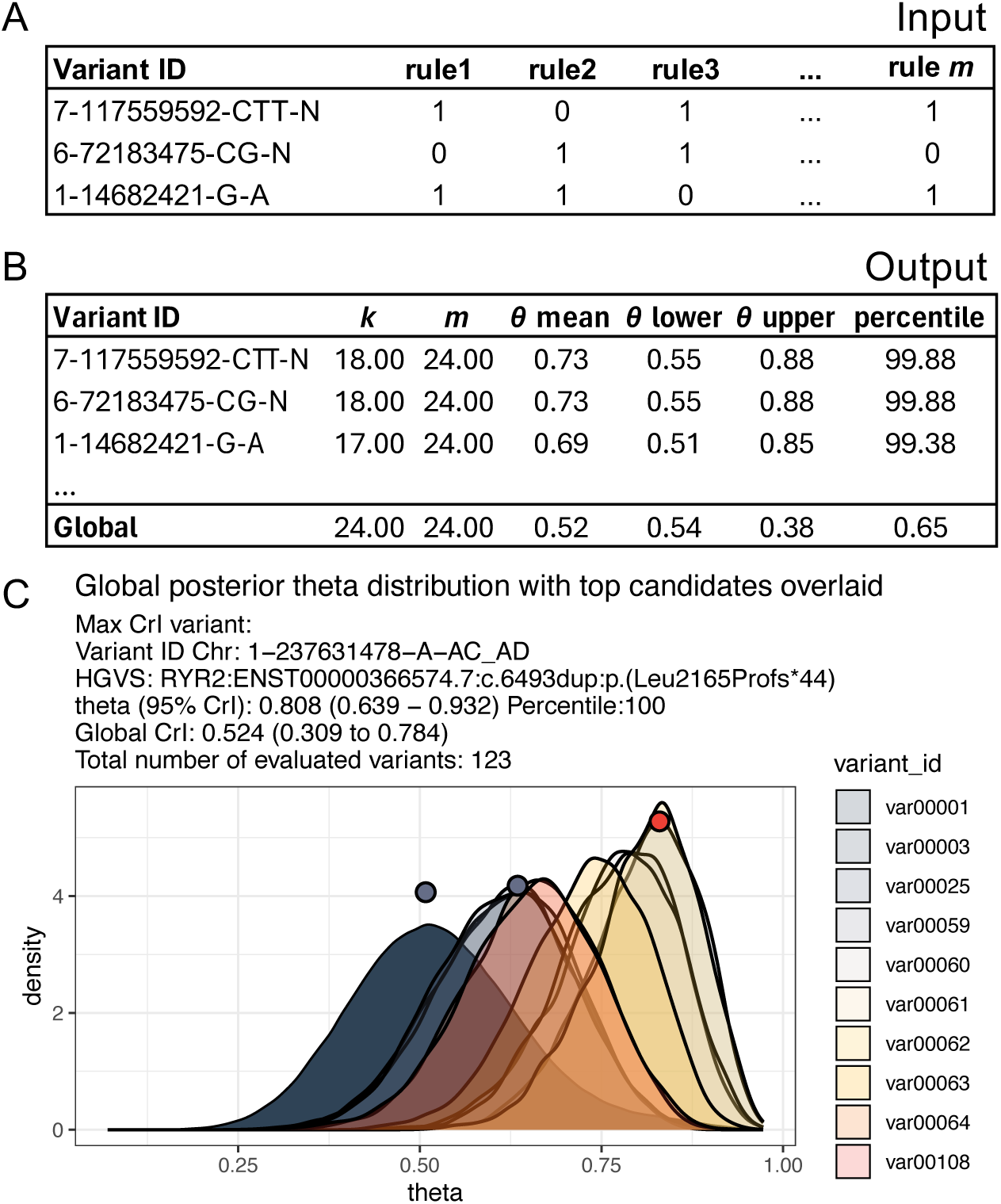
Input, and output, and GIAB WGS trio study. **(A)** Binary evidence matrix with variants as rows and evidence rules as columns, indicating whether each rule is satisfied (1) or not (0). **(B)** QuantBayes output summarising, per variant, the number of satisfied rules *k*, total evaluated rules *m*, posterior mean *θ*, its 95% CrI, and the genome-wide percentile of *θ*. **(C)** Genome-wide mixture distribution of *θ* across all variants (dark), with transparent overlays for top Exomiser-prioritised candidates. The red dot marks the variant with the highest evidence sufficiency CrI, and grey dots indicate the two phenotype-based Exomiser rank 1 predictions. Elevated values reflect stronger interpretative evidence, not causality.

### Reproducibility and compatibility

Input and output formats are compatible with standard variant tables, including decomposed VCF records and PLINK text filesets (23; 24). Variant identifiers may follow the VCF CHROM−POS−REF−ALT convention when the ID field is absent, and outputs preserve input row-order. All computations in Quant ES use deterministic evaluation of the Beta posterior. Where Monte Carlo sampling is used for genome-wide summaries, fixed seeds may be supplied. Evidence standards and interpretation rule sets are provided as registered YAML or JSON specifications (8), ensuring consistent reconstruction of evidence matrices and posterior outputs across sites. Integration with workflow managers such as Snakemake (25) and Nextflow (26) supports reproducible deployment in existing analysis environments.

## 4 Results

### Per-variant summaries

**Figure 1 (B)** shows a basic QuantBayes output. A structured report generated from the same values is provided in **Box 1**. Traditionally, variant interpretation pipelines identify one or more candidate causal variants based on their internal analyses. QuantBayes answers a different question: it quantifies, for every variant, how much verifiable internal and external scientific evidence is available, so the strength of support for any nominated candidate is reported. Using the rule set that defines the evidence flags, QuantBayes provides this transparent and reproducible measure that can be understood and compared across institutions.

##### Box 1: Example of verifiable evidence sufficiency output

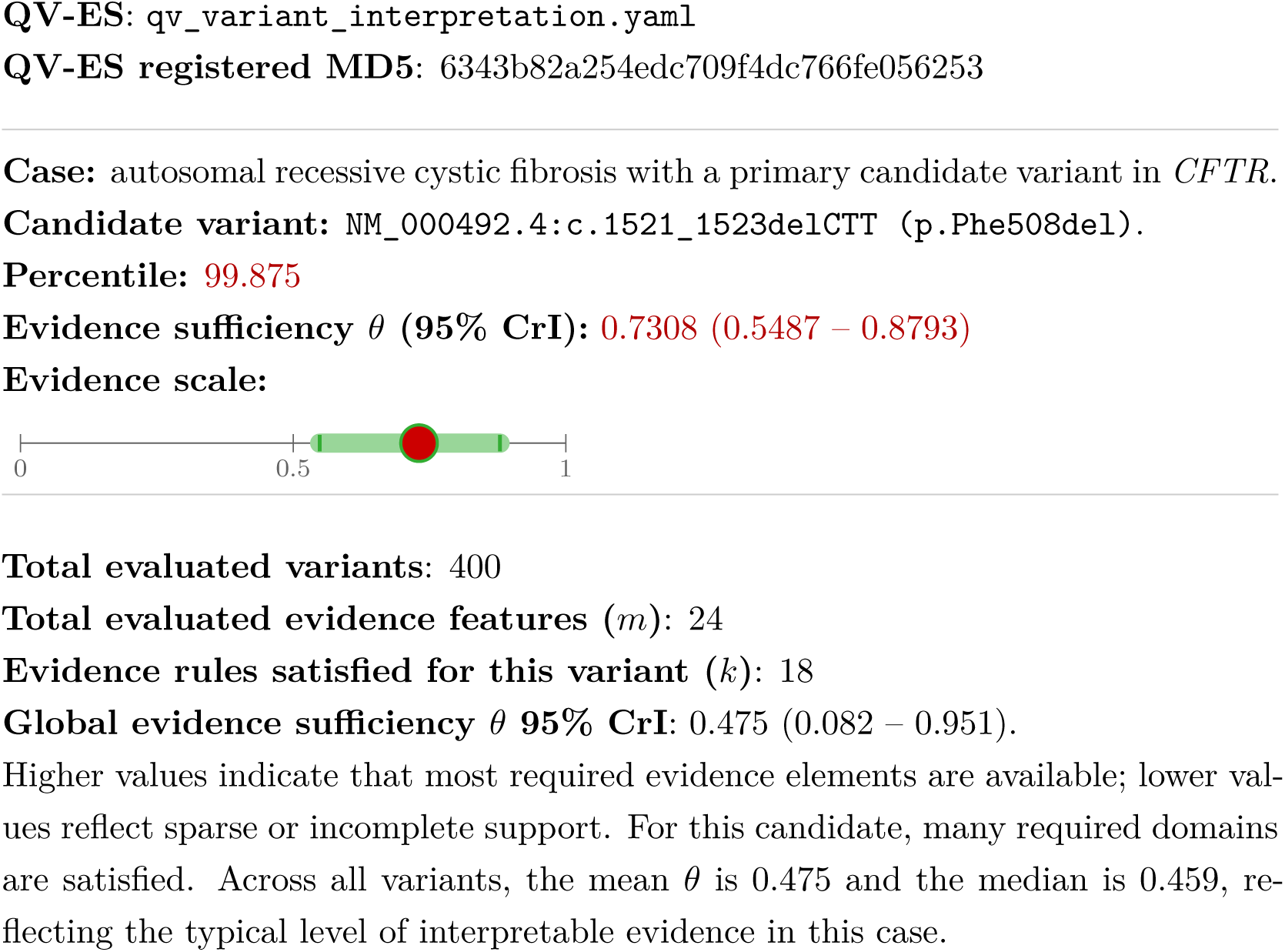

### Validation study evaluation on a public WGS trio

The Quant ES framework was applied to the GIAB Trio (HG005–HG007, GRCh38 v4.2.1) and analysed with Exomiser (19), producing a ranked annotated variant list typical of clinical WGS analysis for Mendelian disease. **Figure S7 - S8** show the evidence standard flag rules before and after interpretation, reflecting the confirmation of evidence features such as population frequency and inheritance consistency.

**Figure 1 (C)** displays the genome-wide mixture distribution of *θ* with overlaid density curves for the top Exomiser candidates. The variant with the strongest verifiable evidence sufficiency was *RYR2*:ENST00000366574.7:c.6493dup:p.(Leu2165Profs*44) with Autosomal Dominant (AD) Mode of Inheritance (MOI), with *θ* (95% CrI) = 0.808 (0.639 – 0.932) and a percentile of 100. This candidate emerged from 123 evaluated variants derived from an initial WGS call set of 5,188,814 variants. Variants rising above the genome-wide mixture are supported by substantially more interpretable evidence. Quant ES therefore quantifies the strength of support for hypotheses proposed by tools such as Exomiser, in a manner directly comparable across approximately one hundred variant effect and prioritisation predictors (11; 19; 27).

Taken together, these results show that Quant ES provides a reproducible, transparent, pipeline-independent measure of evidence availability. The framework separates the evidence layer from provider-specific logic and supports consistent interpretation, verification, and shared use across clinical, research, and commercial settings.

### Performance benchmarks

QuantBayes runs efficiently across both clinical and cohort-scale analyses (**Figure S1**). Runtime scales with the number of variants and evaluated evidence rules. Clinical diagnostic-sized inputs complete in negligible time, while matrices containing up to ten million variants and five hundred evidence sources complete in under ten minutes on a standard laptop. Precompiled C binaries run directly on HPC systems without requiring administrator privileges. The method is therefore practical for routine clinical use and large cohort studies without specialised hardware.

### Public and patient involvement

Public and Patient Involvement and Engagement (PPIE) in genomics is constrained by a persistent practical problem where patients and secondary institutions cannot guarantee which analyses were performed, and results therefore cannot be reused without repeating analysis (28). This arises because evidence checks are embedded within local pipelines rather than recorded in a standard portable, verifiable form. QuantBayes resolves this by running on a pipeline-independent checklist of verifiable evidence that accompanies genomic results. Evidence availability is encoded as a registered, versioned binary matrix derived from explicit rule sets, making clear which evidence domains were evaluated and which were satisfied for each variant. Building on the qualifying variant framework (8), which captured analytic intent and PPIE input as structured metadata, QuantBayes extends this principle by enabling PPIE groups to contribute to the construction of standardised evidence rule sets, supporting transparent verification and reuse of genomic results across institutions (22; 29).

## 5 Conclusion

We introduced a Bayesian framework for quantifying how complete the verifiable scientific evidence is for each genomic variant in Mendelian disease analysis. By operating directly on an evidence matrix and remaining independent of provider specific pipelines, the method delivers a transparent and reproducible evidential layer. A single posterior sufficiency parameter, with uncertainty and genome wide context, summarises both variant level evidence completeness and relative standing among alternatives. This enables consistent interpretation, direct comparison of results across institutions, and reliable reuse of genomic evidence without reanalysis, supporting interoperability in genomic medicine.

## Data Availability

The data used in this manuscript is derived from open sources which are cited in methods. The data generated is available from the Zenodo repository (https://doi.org/10.5281/zenodo.17919369). The resulting data are also reproducible using the code repository. WGS GIAB trio data (HG005-HG007, PRJNA200694, GRCh38 v4.2.1) from Wagner et al.
(18) with the benchmark VCF files for small variants from GRCh38 at https://ftp-trace.ncbi.nlm.nih.gov/ReferenceSamples/giab/release/.

## Acknowledgements

We would like to thank all the patients and families who have been providing advice on SwissPedHealth and its projects, as well as the clinical and research teams at the participating institutions.

## Data and software availability

All data used in this study are derived from open sources referenced in the Methods section. WGS GIAB trio data (HG005–HG007, PRJNA200694, GRCh38 v4.2.1) described by Wagner et al. (18), together with the benchmark small variant VCFs on GRCh38 available at https://ftp-trace.ncbi.nlm.nih.gov/ReferenceSamples/giab/release/. All results are reproducible using the accompanying code repository. Generated data are deposited in the Zenodo archive. The Zenodo archive includes precompiled QuantBayes binaries for macOS (Intel), Linux (x86_64), and R source code https://doi.org/10.5281/zenodo.17919369 (20). The QuantBayes R package is freely available from CRAN https://doi.org/10.32614/CRAN.package.quantbayes (21) and its source repository is at https://github.com/DylanLawless/src-quantbayes_package. All software is provided under MIT license as code intended to be run, modified, or embedded in software. The SGA-QEM-1.0 standard is recommended and is CC BY 4.0 for reuse with attribution. Downstream users may use outputs commercially under MIT or CC BY 4.0 without impact on their own IP or patents, subject only to attribution or licence notice requirements.

## Contributions

DL produced the work and contributed to manuscript writing. AS contributed to the manuscript writing and software testing. LJS contributed to the manuscript writing and funding.

## Ethics statement

The projects were approved by the respective ethics committees of all participating centers (Cantonal Ethics Committee Bern, approval number KEK-029/11) and the study was conducted in accordance with the Declaration of Helsinki.

## Supplemental

### Example QV ES flags and interpretation rules

##### Box S1: Example QV ES YAML (parental genotype completeness)

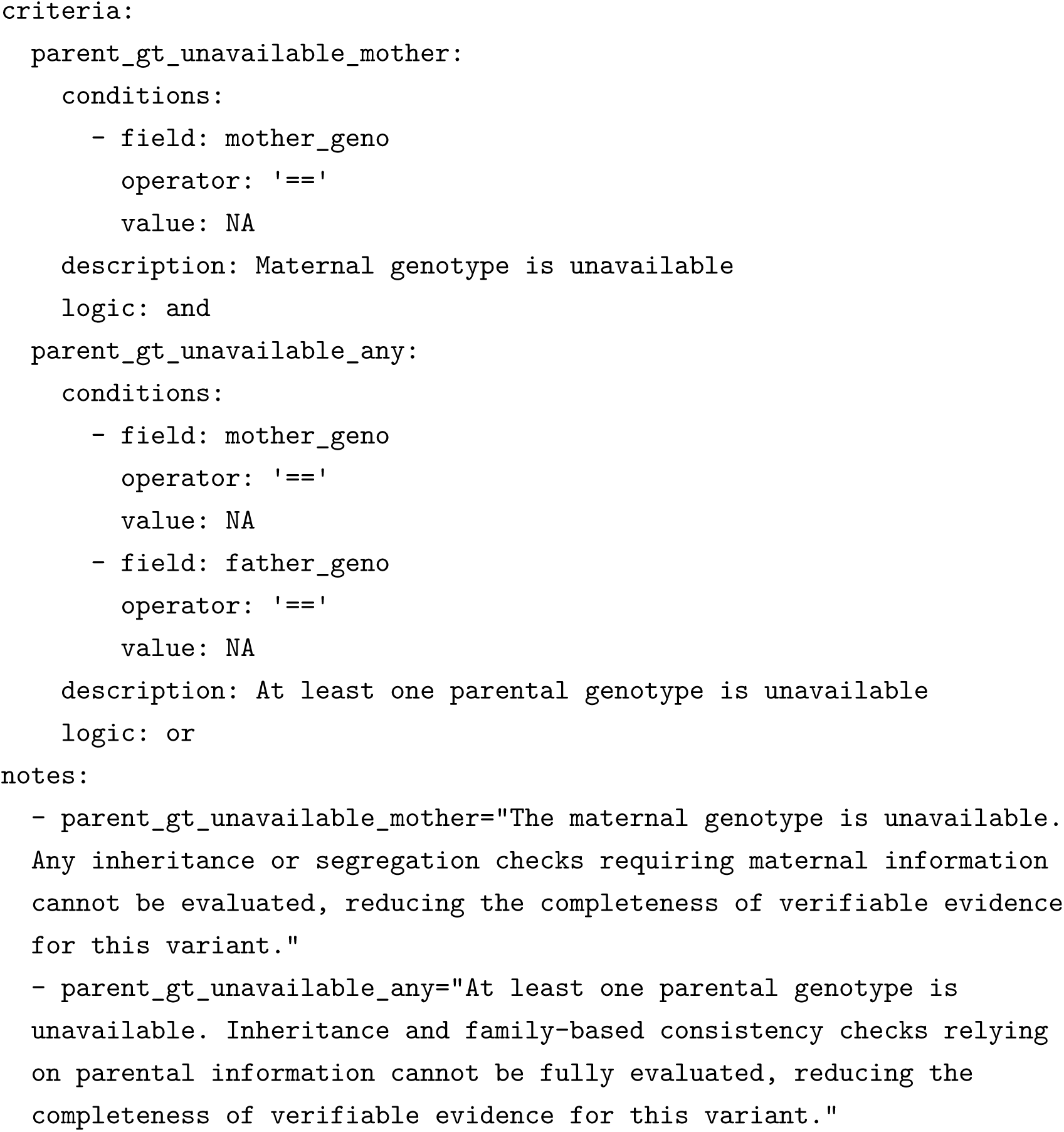

##### Box S2: Example QV ES YAML (gnomAD quality flags)

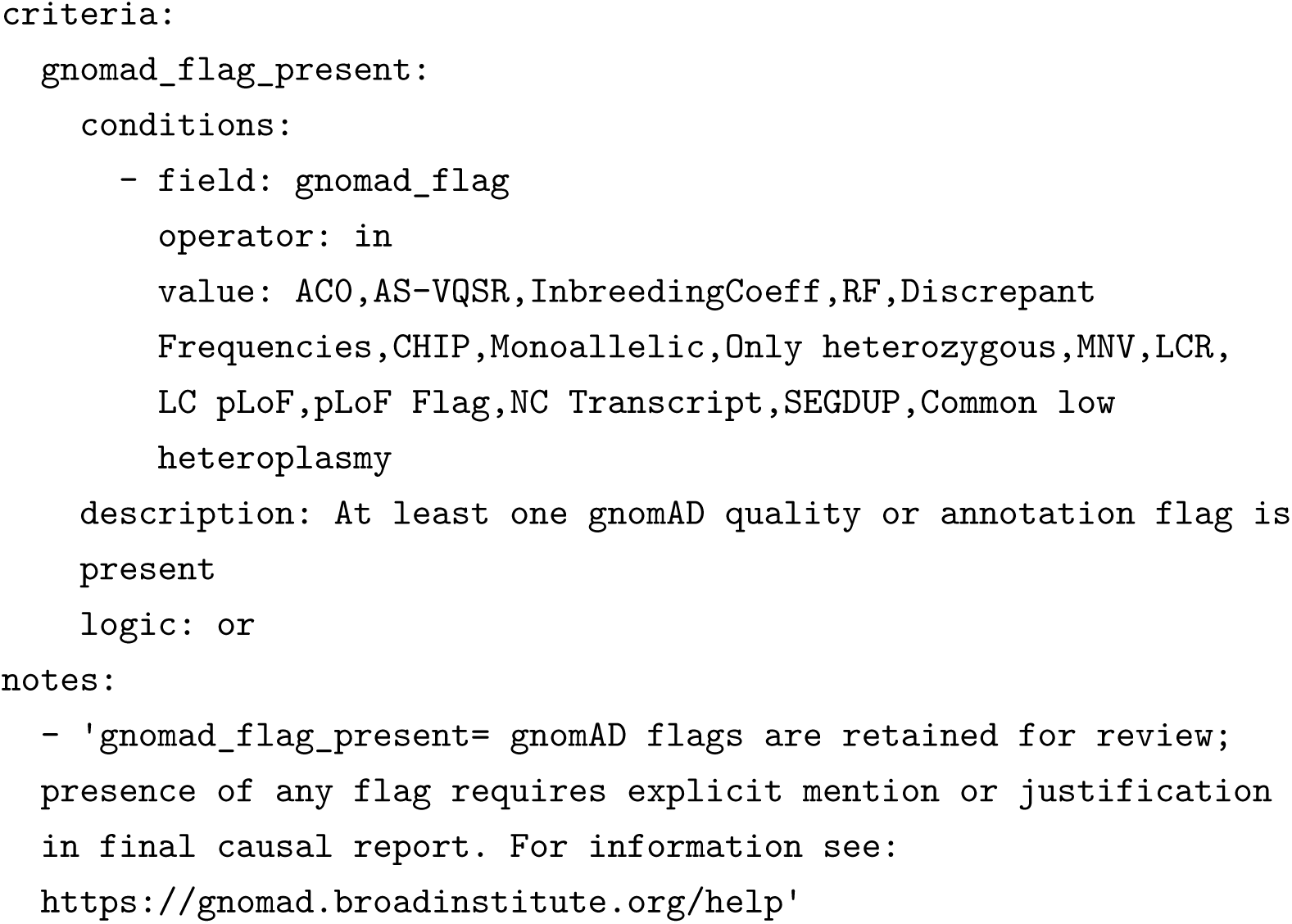

##### Box S3: Example of verifiable evidence sufficiency output

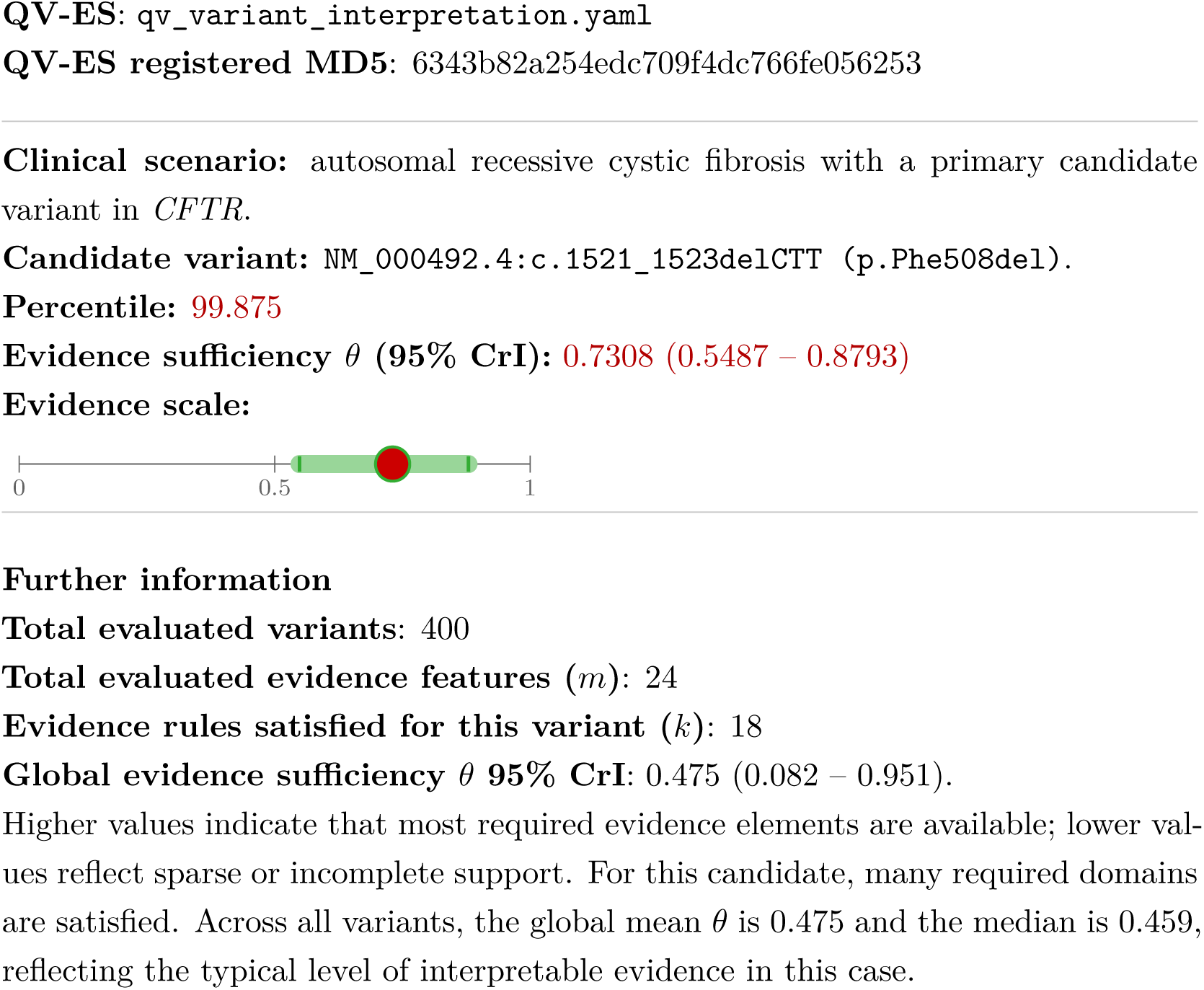

**Figure S1:**
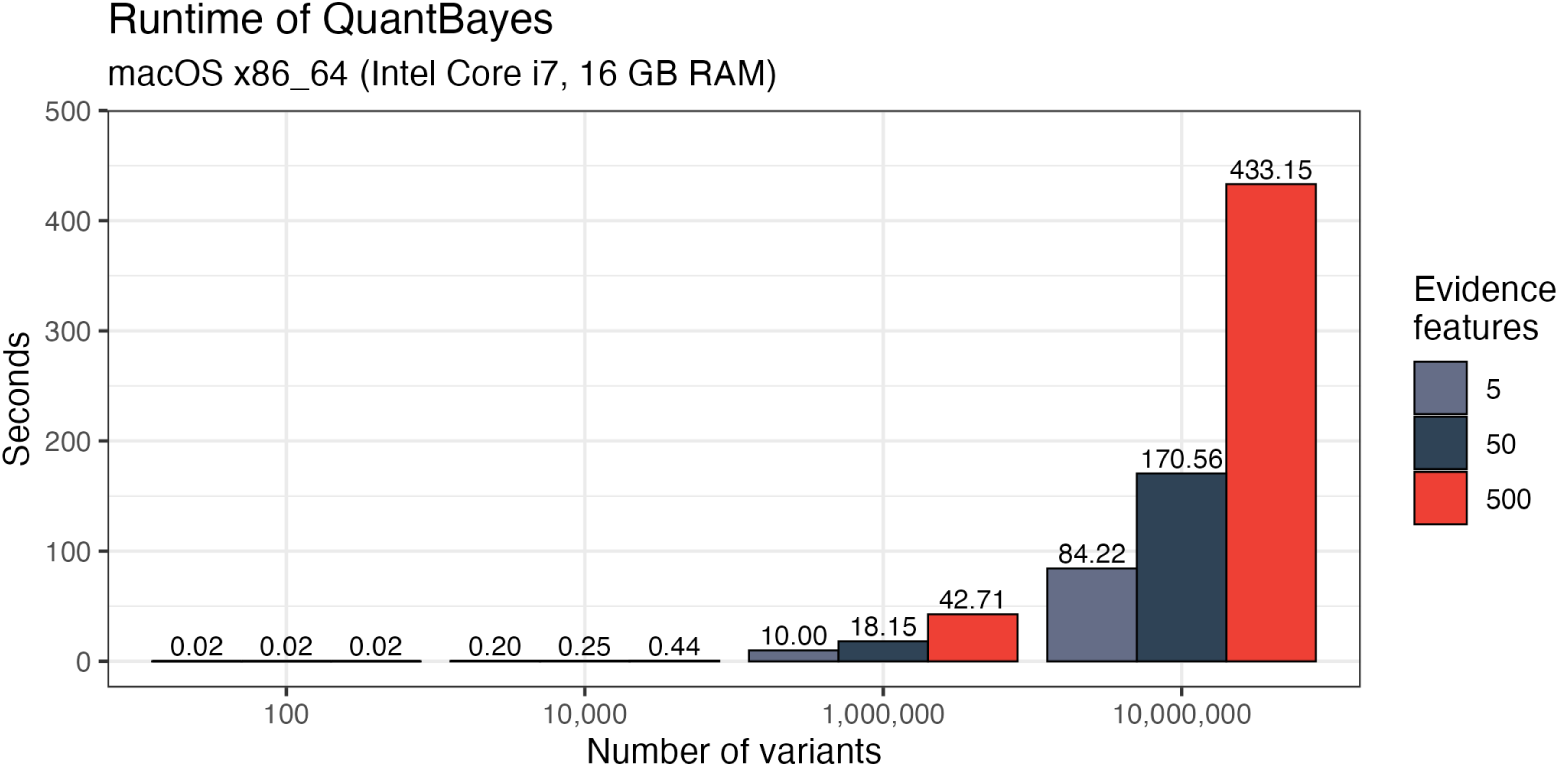
Runtime of QuantBayes for varying numbers of variants (rows) and evidence features (columns). Clinical sized inputs complete in under 0.02 seconds. Inputs up to ten million variants complete in under 10 minutes, demonstrating scalability from routine diagnostics to biobank scale analysis.

**Table S1:** Standard evidence interpretation rules. Each rule maps raw flags (TRUE, FALSE, NA) to binary evidence indicators based on whether the observed data contradict the underlying hypothesis. Rule names describe what a TRUE flag detects, and TRUE preferably corresponds to a contradiction or weakening signal. A value of 1 indicates that no contradiction is observed. Missing data (NA) never counts as evidence present. These rules assess verifiable evidence only; pathogenicity, penetrance, or counterfactual reasoning are handled upstream.

| Rule name | Flag | Meaning | $X_{ij}$ |
| --- | --- | --- | --- |
| parent gt unavailable | TRUE | Parental genotype data unavailable | 0 |
| parent gt unavailable | FALSE | Parental genotype data is available | 1 |
| parent gt unavailable | NA | Availability unknown | 0 |
| parent gt mechanistically inconsistent ar | TRUE | Parental genotype contradicts AR inheritance | 0 |
| parent gt mechanistically inconsistent ar | FALSE | No contradiction with AR inheritance observed | 1 |
| parent gt mechanistically inconsistent ar | NA | Parental genotype unavailable | 0 |
| compound het phase inconsistent | TRUE | Cis or phase contradicts compound heterozygosity | 0 |
| compound het phase inconsistent | FALSE | Trans or phase supports compound heterozygosity | 1 |
| compound het phase inconsistent | NA | Phase information unavailable | 0 |
| denovo mechanism not excluded | TRUE | Parental genotypes exclude a de novo explanation | 0 |
| denovo mechanism not excluded | FALSE | Parental genotypes do not exclude a de novo mechanism | 1 |
| denovo mechanism not excluded | NA | Parental genotypes unavailable | 0 |
| unaffected relative homozygous alt | TRUE | Unaffected parent or close relative carries the same homozygous ALT genotype | 0 |
| unaffected relative homozygous alt | FALSE | No unaffected relative with the same genotype observed | 1 |
| unaffected relative homozygous alt | NA | Phenotype or genotype information unavailable | 0 |

**Table S2:** Required QuantBayes input format. The first column must to contain a unique variant identifier, followed by the rule-based, *m* binary evidence indicators. This structure is required for both the command-line and R implementations. An example evidence feature is whether the proband genotype matches the required MOI.

| variant_id | rule1 | rule2 | rule3 | ... | rule$m$ |
| --- | --- | --- | --- | --- | --- |
| 7-117559592-CTT-N | 1 | 0 | 1 | ... | 1 |
| 6-72183475-CG-N | 0 | 1 | 1 | ... | 0 |
| 1-14682421-G-A | 1 | 1 | 0 | ... | 1 |

**Table S3:** QuantBayes output. For each variant, *k* is the number of evidence rules satisfied, *m* is the total number of evaluated rules, *θ* is the posterior evidence sufficiency, and the 95% CrI and percentile describe its position within the genome wide distribution. The variant id column is shown here for illustration and is carried through from the input matrix; the raw text output omits this column and preserves the original row order. The global summary is reported in a separate_global file.

| variant id | $k$ | $m$ | $\theta$ mean | $\theta$ lower | $\theta$ upper | percentile |
| --- | --- | --- | --- | --- | --- | --- |
| 7-117559592-CTT-N | 18 | 24 | 0.7308 | 0.5487 | 0.8793 | 99.875 |
| 6-72183475-CG-N | 18 | 24 | 0.7308 | 0.5487 | 0.8793 | 99.875 |
| 1-14682421-G-A | 17 | 24 | 0.6923 | 0.5061 | 0.8505 | 99.375 |
| ... | ... | ... | ... | ... | ... | ... |
| Global | 24 | 0.5192 | 0.5385 | 0.3846 | 0.6538 | 0.9500 |

**Figure S2:**
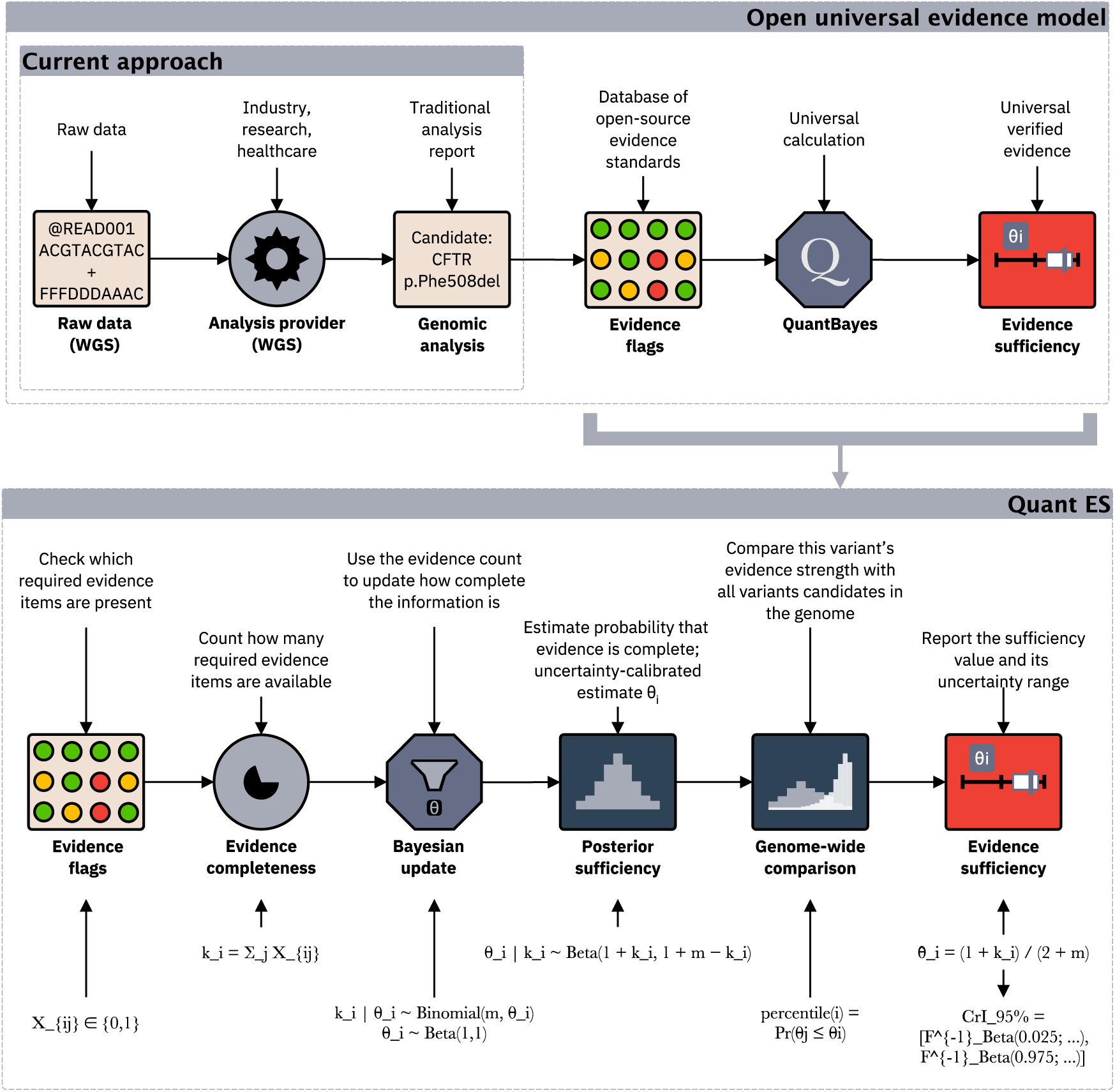
Supplementary Quant ES calculation steps. The schematic defines the symbols used in the Quant ES workflow. The input is a binary matrix *X_ij_* with variants as rows and evidence features as columns. For variant *i*, the evidence count is 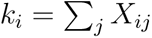 and the number of evaluated features is *m*. The parameter *θ_i_* denotes the evidence sufficiency for variant *i*. The figure shows the posterior *θ_i_* | *k_i_*, its mean, its CrI, and the genome wide percentile Pr(*θ_j_* ≤ *θ_i_*), which are the reported Quant ES outputs.

**Figure S3:**
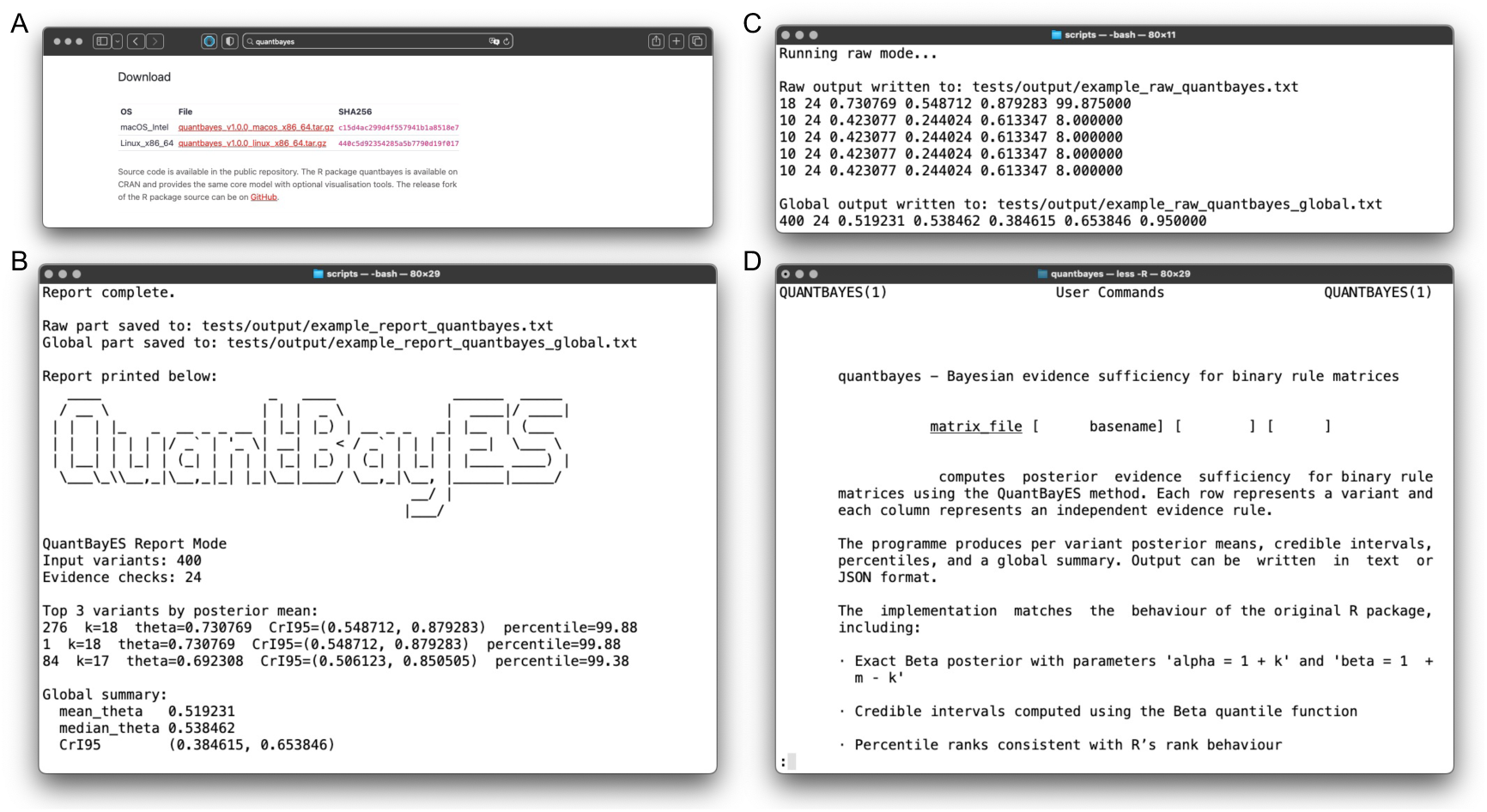
QuantBayes example outputs. (A) The cross-platform R package and executable binaries may downloaded from the public release archive for within research, clinical, or commercial environments. (B) Example console output produced using report mode, summarising top ranked variants and global mixture statistics. (C) Example raw output mode, showing per variant posterior means, CrIs, and percentiles. (D) Extract from the QuantBayes manual page illustrating usage examples.

**Figure S4:**
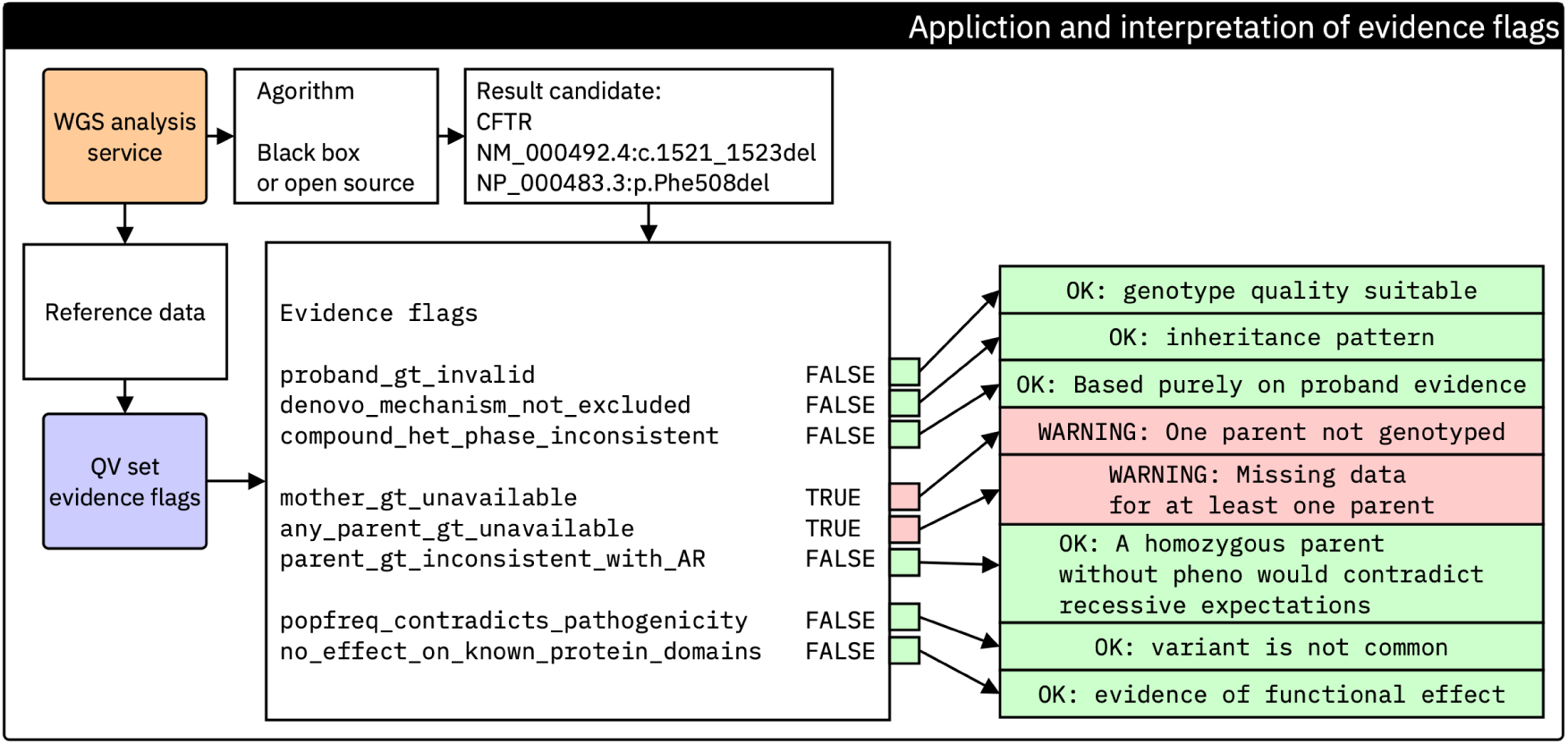
Generation and interpretation of QV ES evidence flags. After secondary analysis, each variant is evaluated against the registered evidence standard flag rule set (SGA-QEM-1.0) (7). Each rule checks whether a required piece of biological or functional evidence is present. The results form a set of raw evidence flags (TRUE, FALSE, or NA).

### Validation figures

**Figure S5:**
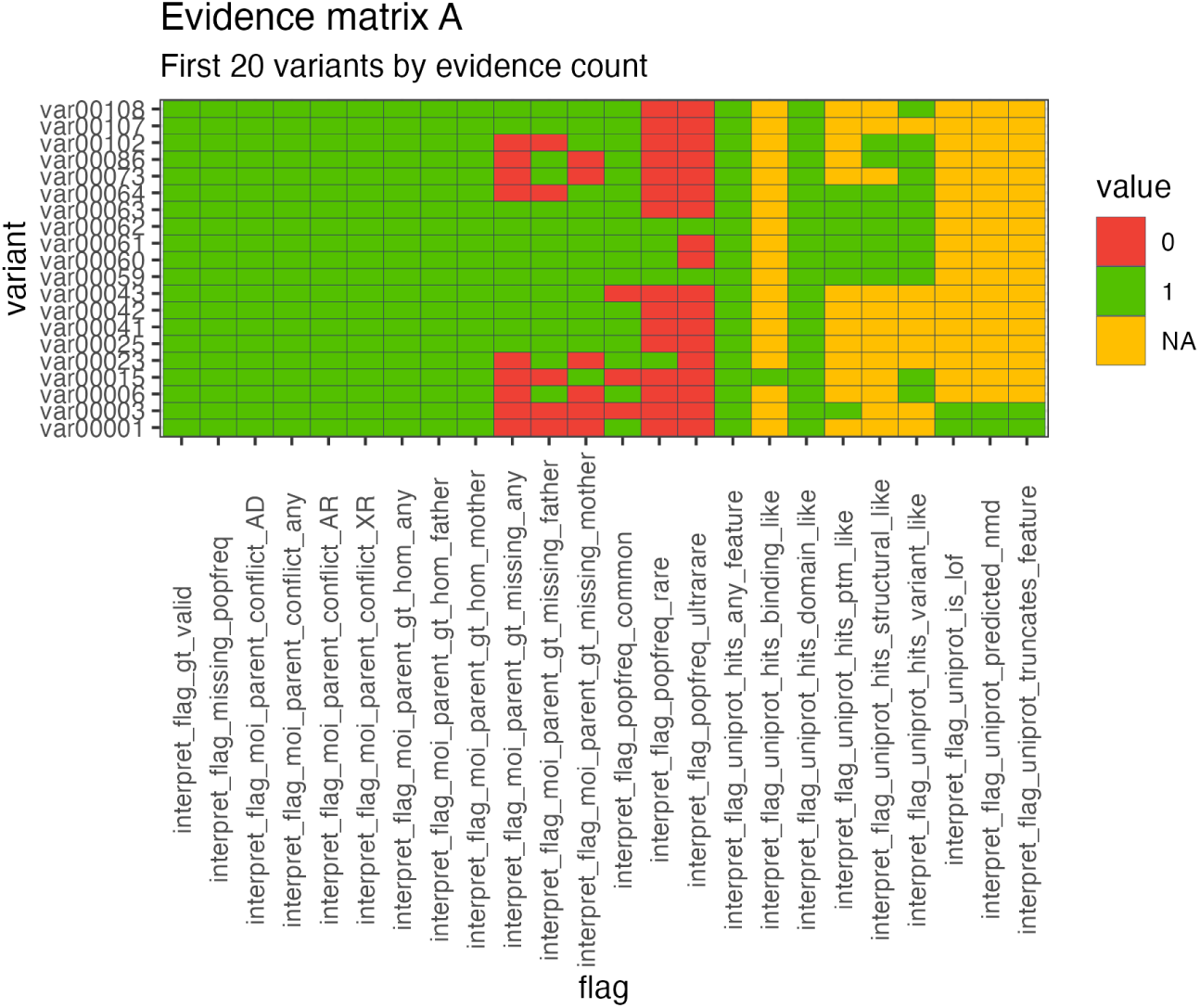
Standard evidence interpretation matrix (GIAB WGS trio). Each row corresponds to a variant and each column corresponds to a evidence standard rule. Cells encode the derived binary evidence outcome (1 = evidence present, 0 = evidence missing) after converting the Exomiser outputs using the flag set. The heterogeneity across rules illustrates the variability in available scientific evidence in a real whole genome analysis. We illustrate NA evidence in yellow indicating that this was specifically checked and no current evidence exists.

**Figure S6:**
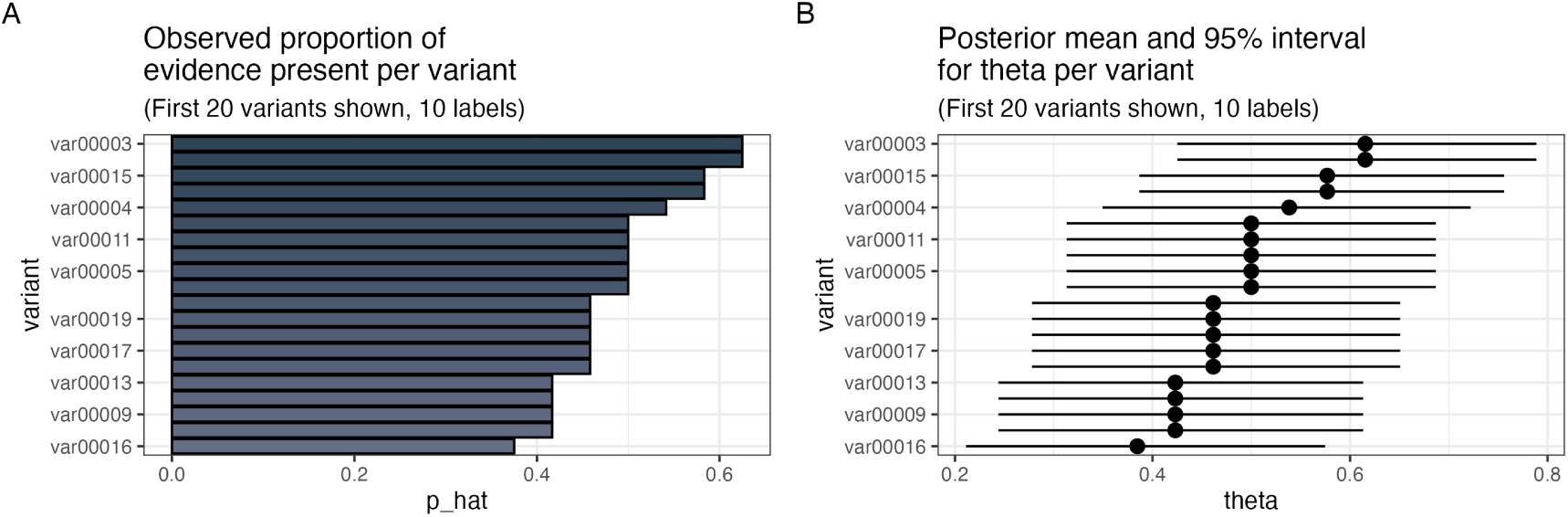
Evidence sufficiency across variants (GIAB WGS trio). **(A)** Observed proportion of satisfied evidence rules. Bars show *p̂_i_* = *k_i_*/*m* for the first 20 variants, illustrating the completeness of biological and functional evidence available for each locus. **(B)** Posterior means and 95% CrIs for *θ_i_*. The Bayesian model converts evidence counts into uncertainty calibrated estimates of evidence sufficiency under real clinical analysis conditions.

Figure S7: **Raw evidence flags from Exomiser outputs (GIAB WGS trio).** Rows represent variants and columns correspond to evidence standard rules. Cells show the direct flag outcomes returned by the evaluation: TRUE (evidence present), FALSE (evidence absent), and NA (evidence available within the dataset to confirm). These raw flags provide a standardised and provider independent summary of the evidence underlying each variant before statistical interpretation.

Figure S8: **Binary evidence interpretation matrix (GIAB WGS trio).** The registered interpretation rules convert the raw flags into a binary matrix suitable for Quant ES analysis. Cells indicate evidence present (1) or evidence missing (0), following the clinical interpretation logic of the evidence standard. The resulting matrix forms the input for Quant ES and enables consistent evidence comparison across all evaluated variants.

